# Percutaneous Peripheral Nerve Stimulation as an Ancillary Treatment Following Nerve Repair Surgery

**DOI:** 10.1101/2025.06.05.25329049

**Authors:** Shyam Maisuria, Antonio Mondríguez-González, Texakalidis Pavlos, Kevin N. Swong, Colin K. Franz

**Affiliations:** Frank H. Netter MD School of Medicine at Quinnipiac University, 370 Bassett Road, North Haven, CT, 06473, USA; Shirley Ryan AbilityLab, 355 E. Erie Street, Chicago, IL, 60611, USA; Department of Physical Medicine and Rehabilitation, Northwestern University Feinberg School of Medicine, Chicago, IL, USA; McGaw Medical Center, Northwestern University Feinberg School of Medicine, 420 E Superior St STE 9-900, Chicago, IL 60611, USA; Department of Neurological Surgery, Northwestern University Feinberg School of Medicine, 676 N. St. Clair, Suite 2210, Chicago, IL, 60611, USA; The Ken and Ruth Davee Department of Neurology, Northwestern University Feinberg School of Medicine, 303 E. Chicago Ave., Chicago, IL, USA; Querrey Simpson Institute for Bioelectronics, Northwestern University, Technological Institute, A/B Wing, 2145 Sheridan Road, Evanston, IL, 60208, USA

**Keywords:** Peripheral nerve stimulation, Neuropathic pain, Nerve repair surgery, Pain management, Axon regeneration

## Abstract

**Background and Aims:** Neuropathic pain after peripheral nerve injury (PNI) severely degrades quality of life. Peripheral nerve stimulation (PN-Stim) offers a potential treatment for pain relief after PNI, although its efficacy in managing neuropathic pain post-nerve repair surgery remains unexplored.

**Methods:** We analyzed 16 consecutive patients aged 18 and above who received PN-Stim implantation post-nerve repair surgery. Patients under 18 years of age, pregnant, and those with PN-Stim for off-label uses were excluded from our study. The primary outcome was pain score reduction, which was evaluated by Visual Analog Scale (VAS). The secondary outcomes included motor functional recovery and opioid usage which were evaluated by manual muscle testing (MMT) and Morphine Milligram Equivalents (MME) quantification, respectively. Statistical analyses utilized paired two-tailed T-tests and Wilcoxon sign tests, contingent on data normalcy.

**Results:** Pain scores decreased significantly post-PN-Stim implantation (mean pre-placement: 7.35, post-placement: 2.56; p < 0.05). MMT scores showed improvement in 13 patients, with two achieving the maximum MMT score (p < 0.05). Chronic opioid usage was observed to decrease in 6 out of 7 patients who were using them (p < 0.05). No significant adverse effects were seen after implantation.

**Interpretation:** The results suggests that PN-Stim is a safe and effective treatment for reducing pain in patients with PNI who have undergone post-nerve repair surgery, without interfering with motor recovery. Future prospective research to explore the role of PN-Stim in pain management and its interaction with functional recovery is warranted.

## Introduction

Conservative estimates indicate that peripheral nerve injuries (PNIs) affect more than 200,000 people annually in the United States (2). Despite peripheral axons having some capacity for regeneration, PNIs frequently result in incomplete functional outcomes. In the most severe cases, PNIs can lead to irreversible muscle paralysis and debilitating neuropathic pain, severe enough to make patients consider limb amputation as a treatment (1, 5). Neuropathic pain is a particularly common and difficult-to-treat consequence of PNIs (2). It frequently presents as severe burning, tingling, or shock-like sensations and can profoundly diminish a patient’s daily functioning and engagement in daily activities. Current treatment options for neuropathic pain are inadequate for many patients (1). For instance, pharmacological therapies have limitations due to their side effects, potential for dependence, and reduced efficacy over time. Thus, there is an urgent need for alternative strategies to manage this condition, particularly in patients recovering from nerve repair surgery.

Peripheral nerve stimulation (PN-Stim) has emerged as a promising approach for managing chronic pain, including neuropathic pain (18). It has been hypothesized that the therapeutic effect of PN-Stim relates to the gate control theory, in which non-painful stimuli can inhibit the transmission of pain signals via A-delta and C-fibers (4). Additionally, PN-Stim may prevent the depolarization of nerve cell membranes, thereby reducing the excitability of pain-transmitting fibers (4). Despite its growing use for pain management in general, the role of PN-Stim in addressing neuropathic pain after PNI and in combination with nerve repair surgery is not defined.

In this study, we explore the effects of PN-Stim on neuropathic pain in patients with severe PNI that have undergone surgical nerve repair by examining changes in pain scores, motor function, and opioid usage. Our hypothesis is that PN-Stim will provide substantial pain relief while also being safe and not interfering with functional recovery.

## Materials and Methods

### Study Design and Subjects

This study is a retrospective chart review of 16 consecutive adult patients who underwent intraoperative implantation of a commercial percutaneous PN-Stim device (SPRINT PNS System) at the time of surgical nerve repair (Table 1). Sixten patients were identified from an interdisciplinary complex nerve injury clinic at an academic rehabilitation hospital (Shirley Ryan AbilityLab, Chicago, IL, USA). All surgeries were performed by a neurosurgeon (K.N.S.) at an affiliated academic tertiary care hospital (Northwestern Memorial Hospital, Chicago, IL, USA). Clinical diagnosis of neuropathic pain was the indication for implantation of the PN-Stim device in all patients. The PN-Stim device is designed for temporary use (60 days), and then the leads were removed by provider using traction. In all cases the device leads were secured at or proximal to the site of nerve repair (Table 1). The study was approved by the Institutional Review Board of Northwestern University.

**Table 1:** Patient Demographics, Injury Characteristics, and Surgical Interventions. This table summarizes the demographic information, injury mechanisms, specific nerve injury sites, the location of percutaneous peripheral nerve stimulator (PN-Stim) placement, and the surgical procedures performed for each patient. Surgical procedures detail the nerve repair (decompression, neurolysis, graft, transfer, and/or resection) that occurred concurrently with or prior to PN-Stim implantation. F/U, Follow-up; PN-Stim, Peripheral Nerve Stimulation.

| # | Age (Sex) | Race/Ethnic. | Injury Mechanism | Nerve Injury Site | PN Stim Site | Surgical Procedure | F/U (days) |
| --- | --- | --- | --- | --- | --- | --- | --- |
| 1 | 70s (M) | White | Compression | Right Ulnar | Right Elbow | Ulnar nerve decompression + PN Stim placement at the elbow | 246 |
| 2 | 60s (M) | White | Stabbing | Left Brachial Plexus & Ulnar | Left Elbow | Resection of ulnar neuroma and sural graft repair + PN Sprint. Also, neurolysis of brachial plexus | 146 |
| 3 | 30s (M) | White | Laceration | Right Median & Ulnar. | Right Elbow. | Resection of ulnar neuroma and sural graft repair + PN Sprint. Also, neurolysis of median. | 561 |
| 4 | 60s (M) | White | Compression | Left Median and Musculocutaneous | Left Forearm | Brachialis-to-anterior interosseus nerve transfer + PN Stim. Also, neurolysis of musculocutaneous and median in arm. | 71 |
| 5 | 30s (F) | White | Compression | Left Radial | Left Arm | Radial nerve neurolysis + PN Stim. | 230 |
| 6 | 30s (F) | White | Motor Vehicle Crash | Left Brachial Plexus | Left Shoulder | Spinal accessory to suprascapular nerve transfer + PN Stim. Also, radial to axillary, ulnar to brachialis transfers. | 57 |
| 7 | 30s (M) | Black | Motor Vehicle Crash | Right Brachial Plexus | Right Shoulder | Sural graft of C5 to suprascapular nerve + PN Stim. | 382 |
| 8 | 20s (M) | Black | Gunshot Injury | Right Sciatic | Left Thigh | Resection of peroneal neuroma and sural graft + PN Stim. Also, neurolysis of sciatic and tibial. | 100 |
| 9 | 20s (M) | Black | Motor Vehicle Crash | Right Brachial Plexus | Right Shoulder | Spinal accessory to suprascapular nerve transfer + PN Stim. Also, radial to axillary, ulnar to brachialis transfers. | 370 |
| 10 | 60s (M) | White | Iatrogenic (Cervical Spine Surgery) | Right C5 Nerve Root | Right Shoulder | Radial-to-axillary nerve transfer + PN Stim. | 373 |
| 11 | 50s (F) | White | Iatrogenic (Hip Surgery) | Left Femoral Nerve | Left Thigh | Obturator to femoral nerve transfer + PN Stim. | 365 |
| 12 | 30s (M) | White | Motor Vehicle Crash | Right Brachial Plexus | Right Shoulder | Spinal accessory to suprascapular nerve transfer + PN Stim. Also, ulnar to brachialis transfers | 179 |
| 13 | 60s (M) | White | Iatrogenic (Cervical Spine Surgery) | Right C5 Nerve Root | Right Shoulder | Radial-to-axillary nerve transfer + PN Stim. Also, ulnar neurolysis at elbow. | 57 |
| 14 | 70s (M) | White | Compression | Left Peroneal Nerve | Left Fibular Head | Peroneal nerve neurolysis at fibular head + PN Stim. | 57 |
| 15 | 60s (F) | Hispanic | Compression | Left Peroneal Nerve | Left Fibular Head | Peroneal nerve neurolysis at fibular head + PN Stim. | 152 |
| 16 | 60s (M) | White | Compression | Right Ulnar Nerve | Right Elbow | Ulnar nerve decompression at elbow + PN Stim. | 202 |

### Study Outcomes

The primary outcome of this study was the Visual Analog Scale (VAS) pain score following PN-Stim implantation with nerve repair surgery. Pain reduction was assessed by comparing the patients’ worst pain scores reported in clinic prior to PN-Stim placement with their worst pain scores after PN-Stim placement. Secondary outcomes included change in manual muscle testing (MMT) scores, and changes in opioid usage, measured in Morphine Milligram Equivalents (MME) before and after PN-Stim implantation. These were analyzed to determine if there was any detrimental impacts due to PN-Stim.

### Data Collection

Pain scores were collected using the VAS (ranging from 1 to 10) and in instances where the VAS was not available in the chart (n=7), the Disabilities of the Arm, Shoulder, and Hand (DASH) questionnaire was used instead, which uses a Likert scale from 1 to 5. To maintain consistency across data sources, DASH scores were categorized as “no pain,” “light pain,” “moderate pain,” and “severe pain,” and then converted to a 1 to 10 scale for comparison with the VAS scores. Pain scores were collected before and after PN-Stim implantation at multiple follow-up medical, physical therapy, and occupational therapy appointments for these patients, and these values were compared to assess pain reduction in the patient population.

To assess opioid usage, prescription records were reviewed to document the use of opioids before and after PN-Stim placement at follow-up appointments with physicians, physical therapy, and occupational therapy. The dates of prescription refills were used to determine if patients were still using opioids, and MME were calculated for those who were prescribed opioids. The MME values from before and after PN-Stim placement were compared to evaluate changes in opioid consumption following the procedure at patient follow-up appointments. To minimize the potential effect of the surgical procedure itself in the pain experienced, only chronic opioid use, and not those prescribed for acute post-operative pain, were considered in this analysis.

MMT scores were retrieved from documentation, either by physicians or physical therapists before and after PN-Stim treatment. MMT scores were categorized into six levels: “no movement,” “flicker,” “severely reduced,” “moderately reduced,” “mildly reduced,” and “normal movement.” Each category was assigned a corresponding numerical value, ranging from 0 for “no movement” to 5 for “normal movement.” These numerical scores were used to assess and compare motor function before and after intervention.

### Statistical Analysis

Due to the sample size of 16 patients, the normality of the data could not be assumed. Instead, kurtosis was used to assess the normality of pain scores, MMT scores, and MME values. Based on the distribution of the data, both parametric and non-parametric tests were employed. A two-tailed paired T-test was used to compare the mean pain, MMT, and MME scores before and after PN-Stim placement. For data that was not normally distributed (MME), the Wilcoxon signed-rank test was used as an alternative. The significance level (alpha) was set at 0.05 for all comparisons. The dataset was assessed for normality, revealing that both pain scores and MMT scores followed a normal distribution, with kurtosis values between –2 and 2, while the MME data displayed a non-normal distribution. For the pain and MMT scores, the data was reported as a mean +/-the standard deviation. For MME, the data was reported as a decrease from the patients’ original MME before the PN-Stim implant.

## Results

Pain scores before and after PN-Stim placement are shown in Figure 1. All 16 patients experienced a reduction in pain following PN-Stim implantation (Figure 1a). The mean pain score before PN-Stim placement was 7.38 +/-2.09, which decreased to 2.56 +/-2.09 after the procedure demonstrating a statistically significant reduction (p < 0.05) (Figure 1b). Importantly, none of the patients experienced adverse effects or elected to have the implant removed before the full treatment period was completed (60 days).

**Figure 1.**
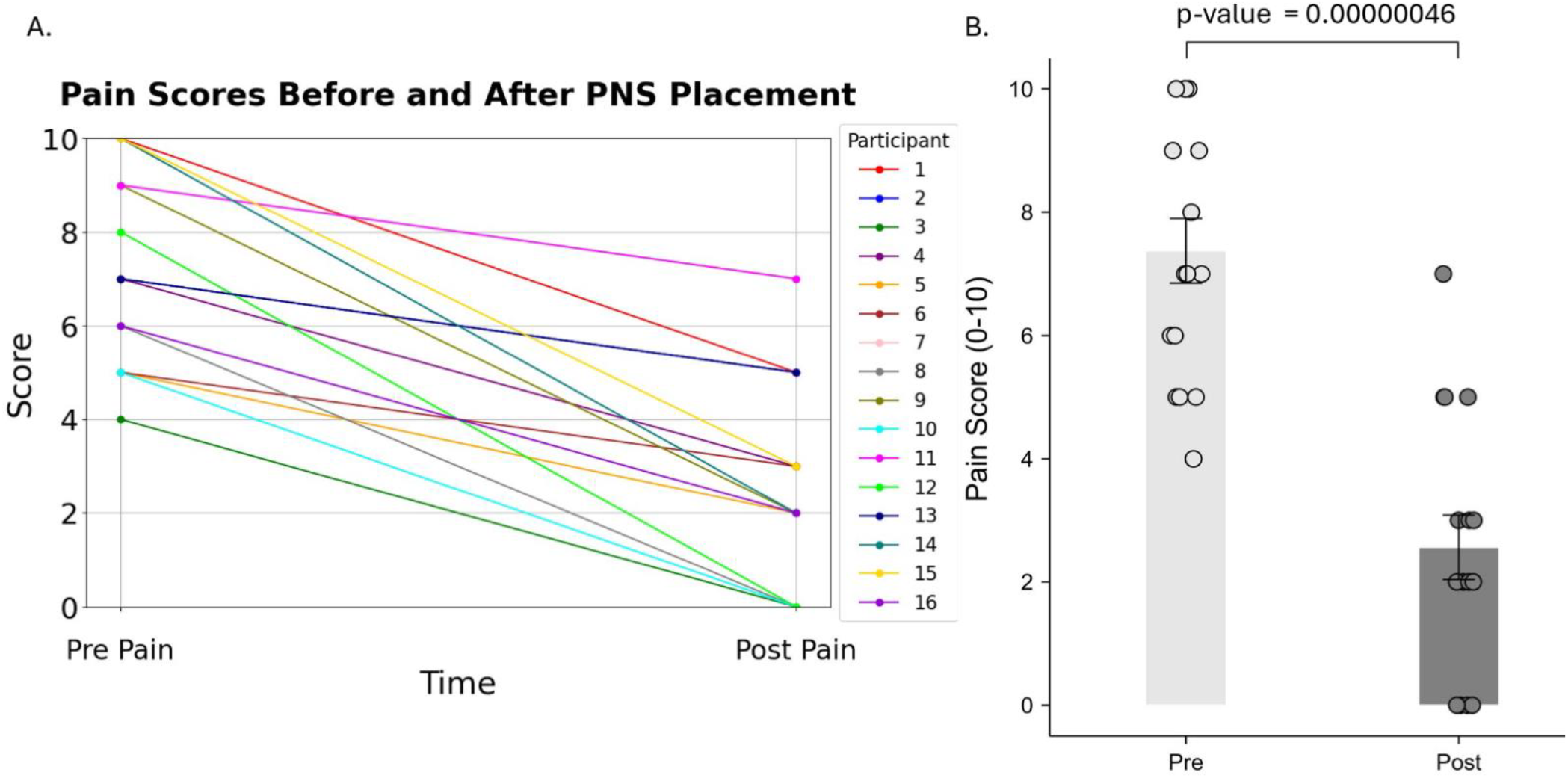
Effect of Nerve Repair and PN-Stim Placement on Pain Scores. (A) Individual pain scores of 16 patients before and after nerve repair and PN-Stim placement, measured using the Visual Analog Scale (VAS) from 1 to 10, where higher scores indicate greater pain intensity. (B) Average pain scores across all patients before and after nerve repair and PN-Stim placement, demonstrating a significant reduction in pain intensity post-treatment.

Opioid usage data revealed that seven patients were prescribed opioids before their nerve repair surgery. Following intervention, six patients reduced their opioid consumption to 0 MME, while one patient showed no reduction in opioid use. Data analysis revealed a statistically significant reduction in opioid use (p < 0.05), as depicted in Figure 2.

**Figure 2:**
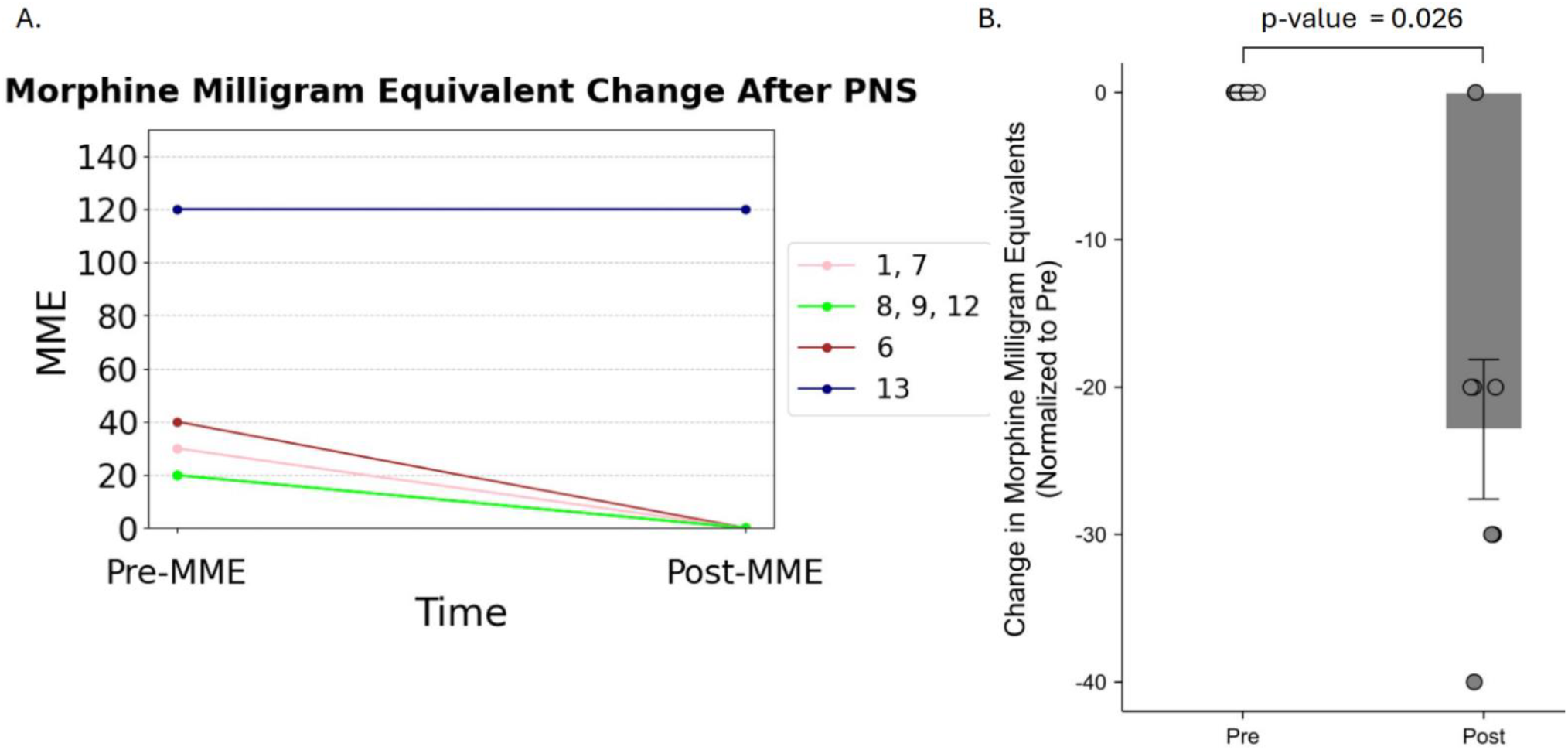
Changes in Morphine Milligram Equivalents (MME) Post Nerve Repair and PN-Stim Placement. (A) Morphine Milligram Equivalents of 7 patients before and after nerve repair and PN-Stim placement, where higher scores indicate greater levels of required pain control. (B) Average MME changes before and after nerve repair and PN-Stim placement, demonstrating a significant reduction in pain medication utilization.

Before PN-Stim placement, four patients demonstrated “no movement”, one patient showed a “flicker,” seven patients showed “severely reduced movement,” three patients had “moderately reduced movement,” and two patients showed “mildly reduced movement.” After PN-Stim placement, three patients showed no improvement, five patients improved by one category, four improved by two categories, two improved by three categories, one patient improved by four categories, and one patient improved by five categories. The comparison of MMT scores before and after PN-Stim intervention through a paired two-tailed T-test demonstrated a statistically significant improvement (p <0.05), as shown in Figure 3B.

**Figure 3:**
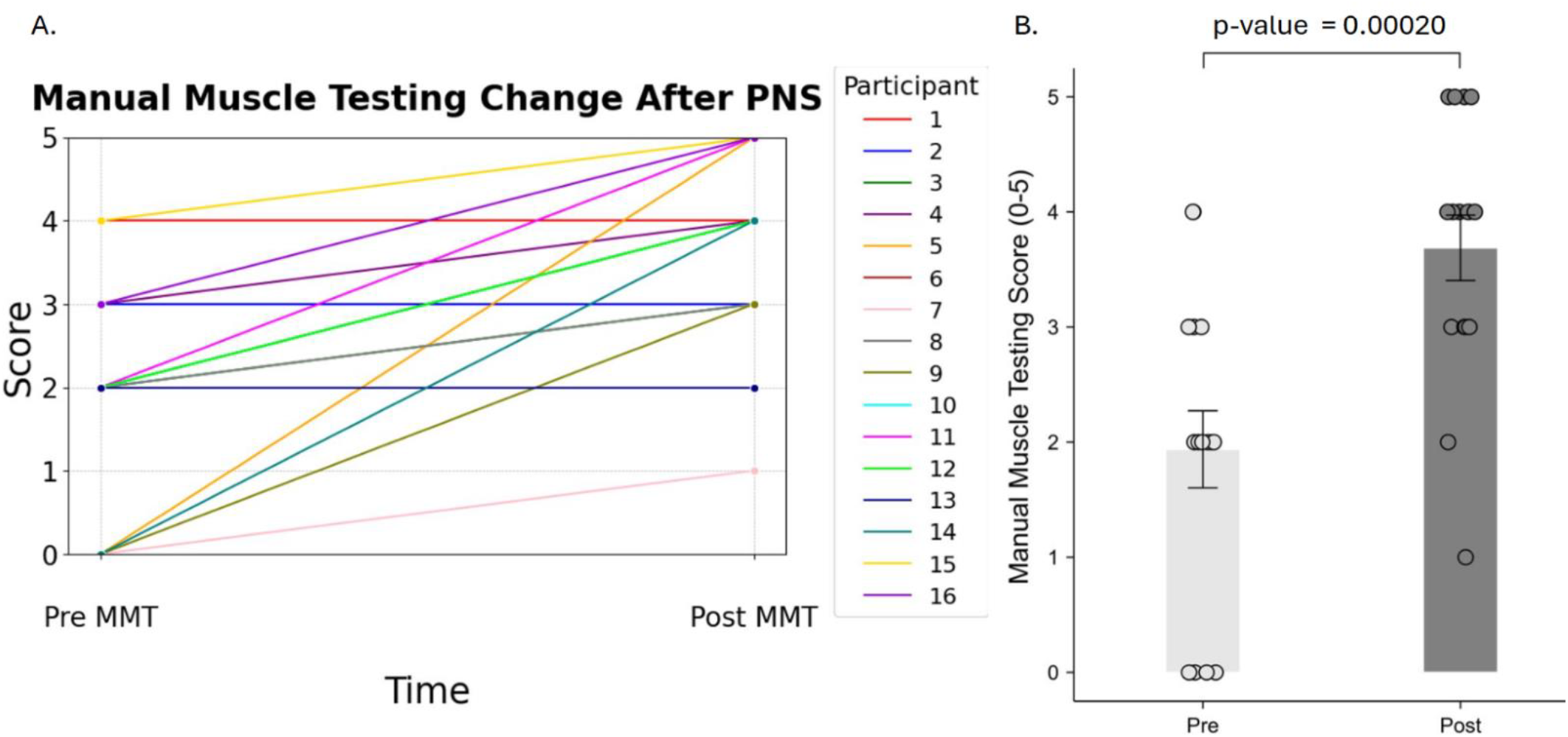
Manual Muscle Testing Change After Nerve Repair and PN-Stim Placement. (A) Individual Manual Muscle Testing Score of 16 patients before and after nerve repair and PN-Stim placement, measured using physical exams performed by providers, where higher scores indicate greater muscle strength. (B) Average MMT scores across all patients before and after nerve repair and PN-Stim placement, demonstrating a significant increase in muscle movement.

PN-Stim significantly reduced pain for patients following nerve repair surgery. However, its efficacy varied between patients. Interestingly, two patients experienced total pain relief while the PN-Stim device was in place, but their pain returned after the implant was removed. Another patient stated that she felt differences in pain when the implant was “on (better)”, vs “off”. This suggests that the inhibition of pain fibers may not persist post-stimulation for some patients. It’s important to note that none of the patients who underwent PN-Stim had a worsened level of pain during or after the course of treatment was finished relative to their baseline pain level

## Discussion

Our study indicates that PN-Stim implantation in conjunction with nerve repair surgery can lead to significant reductions in pain and opioid consumption in people with PNI, while also not interfering with the expected improvements in motor function post-nerve repair. The significant decrease in VAS pain scores and opioid usage, along with gains in MMT strength scores, underscores the potential of PN-Stim as an adjunct to post-surgical rehabilitation. These findings align with prior literature examining the role of PN-Stim in pain modulation for other causes of pain, yet this study adds unique insights into both the feasibility and benefits specifically following nerve repair surgery.

A key finding of this study is the objective demonstration of pain reduction across all patients (Figure 1), with none reporting worsened pain during or after treatment.

Notably, six of the seven patients prescribed opioids prior to PN-Stim intervention discontinued opioid use entirely (Figure 2). We chose to focus on opioid reduction in this study since they have been show significantly more detrimental side effects than other analgesics (3). This reduction in opioid dependence has significant clinical implications, potentially mitigating risks associated with long-term opioid use, such as addiction, respiratory depression, and gastrointestinal side effects (3). While opioid use naturally decreases following surgical recovery, the statistically significant reduction suggests that PN-Stim may accelerate this process or enhance pain control beyond typical recovery trajectories.

Regarding changes in patients’ MMT scores, 13 patients showed motor function recovery in at least one category (Figure 2). In fact, one patient with a femoral nerve transfer and no prior movement before PN-Stim implantation regained full movement after the conclusion of treatment (Patient 11). It’s worth mentioning that none of the patients’ MMT scores worsened after PN-Stim, reassuring us that PN-Stim does not have a detrimental effect on motor function or nerve integrity, thus supporting its safety in those undergoing nerve repair. Future studies incorporating electrophysiological assessments or imaging could provide greater clarity regarding the effects on motor recovery. Notably, acute post-operative electrical stimulation has actually been demonstrated to improve peripheral axon regeneration in preclinical and several single-center clinical trials recently reviewed by Hardy et al. (2024).

Our findings are consistent with previous studies that have examined the effects of PN-Stim on pain. For instance, one case series consisting of patients with chronic lower back pain conducted by Gilmore and colleagues demonstrated a significant reduction in patients’ pain scores one year after PN-Stim treatment (3). The reduction in opioid usage is also consistent with a previous study by Ilfeld and colleagues where they demonstrated that percutaneous nerve stimulation was able to reduce the level of analgesia required following knee-repair surgery (14). This reduction in opioid dependence is particularly significant in light of the ongoing opioid crisis, emphasizing PN-Stim’s potential to offer a safer, non-pharmacologic alternative for post-operative pain management.

Study limitations include the small sample size (n = 16) and reliance on subjective measures such as VAS and MMT scores. Although every effort was made to standardize the collection of these data, inherent variability in patient reporting and clinician assessment introduces the potential for bias. Another limitation is the inability to isolate the effects of PN-Stim from the nerve repair surgery itself, as all patients underwent both interventions. This makes it challenging to determine the extent to which observed improvements are driven by either procedure. Additionally, the absence of long-term follow-up data is a notable limitation. While all patients demonstrated pain relief during the treatment period, the durability of this effect post-implant removal remains uncertain. One patient reported the return of pain following device removal, highlighting the need to investigate whether prolonged or repeated stimulation may sustain pain relief and motor improvements. Future research should explore the optimal duration of PN-Stim treatment and assess outcomes at extended intervals post-treatment.

Mechanistically, while prior studies suggest that electrical stimulation may promote axonal regeneration through the upregulation of neurotrophic factors like brain - derived neurotrophic factor (BDNF) and its downstream signaling pathway (19), our study was not designed to assess these biological factors. Thus, while the observed motor functional improvements are encouraging, they should be interpreted within the context of clinical outcomes rather than underlying cellular mechanisms. Further investigations incorporating animal models and/or new clinical biomarkers (imaging, serum BDNF, etc.) could help elucidate the potential regenerative effects of PN-Stim at a mechanistic level.

In conclusion, PN-Stim shows significant potential as an adjunct therapy for managing neuropathic pain and supporting overall recovery following nerve repair surgery. The observed reductions in pain and opioid use, coupled with the observed improvements in motor function, highlight its promise as a non-pharmacologic ancillary treatment to nerve repair with good safety. By addressing pain, regardless of whether it could theoretically improve neurological recovery, PN-Stim may contribute to reducing opioid use post-operatively. Future studies should focus on elucidating its role in axonal regeneration, optimizing treatment protocols, and expanding its application to broader patient populations to enhance rehabilitation outcomes for individuals recovering from PNI. This should include large randomized controlled trials to confirm these results and determine the long-term effects of this intervention.

## Data Availability

All data produced in the present work are contained in the manuscript

## Acknowledgements

SM was funded by the Shirley Ryan AbilityLab Externship Program for medical students. CKF would like to acknowledge support from the Belle Carnell Regenerative Neurorehabilitation Fund.

## References

1. Smith JK, Myers KP, Holloway RG, Landau ME. Ethical considerations in elective amputation after traumatic peripheral nerve injuries. Neurol Clin Pract. 2014;4(4):280–286. doi:10.1212/CPJ.0000000000000049. PMID: 25279253; PMCID: PMC4160445.

2. Vadivelu N, Kai AM, Kodumudi G, Babayan K, Fontes M, Burg MM. Pain and psychology-a reciprocal relationship. Ochsner J. 2017;17(2):173-180. PMID: 28638291; PMCID: PMC5472077.

3. Gilmore CA, Kapural L, McGee MJ, Boggs JW. Percutaneous peripheral nerve stimulation for chronic low back pain: prospective case series with 1 year of sustained relief following short-term implant. Pain Pract. 2020;20(3):310–320. doi:10.1111/papr.12856. PMID: 31693791; PMCID: PMC7079182.

4. Ong Sio LC, Hom B, Garg S, Abd-Elsayed A. Mechanism of action of peripheral nerve stimulation for chronic pain: a narrative review. Int J Mol Sci. 2023;24(5):4540. doi:10.3390/ijms24054540. PMID: 36901970; PMCID: PMC10003676.

5. Finnerup NB, Kuner R, Jensen TS. Neuropathic pain: from mechanisms to treatment. Physiol Rev. 2021;101(1):259–301. doi:10.1152/physrev.00045.2019. PMID: 32584191.

6. Boyd JG, Gordon T. Neurotrophic factors and their receptors in axonal regeneration and functional recovery after peripheral nerve injury. Mol Neurobiol. 2003;27(3):277–324. doi:10.1385/MN:27:3:277. PMID: 12845152.

7. Chan KM, Curran MWT, Gordon T. The use of brief post-surgical low frequency electrical stimulation to enhance nerve regeneration in clinical practice. J Physiol. 2016;594:3553–3559. doi:10.1113/JP270892.

8. Höke A. Mechanisms of disease: what factors limit the success of peripheral nerve regeneration in humans? Nat Rev Neurol. 2006;2:448–454.

9. Gordon T. The role of neurotrophic factors in nerve regeneration. Neurosurg Focus. 2009;26(2). doi:10.3171/FOC.2009.26.2.E3. PMID: 19228105.

10. Sellinger J. Chronic pain: a biopsychosocial approach. Quinnipiac University Lecture, Yale School of Medicine, Department of Psychiatry.

11. Lu C, Wang Y, Yang S, et al. Bioactive self-assembling peptide hydrogels functionalized with brain-derived neurotrophic factor and nerve growth factor mimicking peptides synergistically promote peripheral nerve regeneration. ACS Biomater Sci Eng. 2018;4(8):2994–3005. doi:10.1021/acsbiomaterials.8b00536.

12. Pandey S, Mudgal J. A review on the role of endogenous neurotrophins and Schwann cells in axonal regeneration. J Neuroimmune Pharmacol. 2022;17:398–408. doi:10.1007/s11481-021-10034-3.

13. Kandel ER, Schwartz JH, Jessell TM. Principles of Neural Science. 4th ed. New York, NY: McGraw-Hill; 2000:482-486. ISBN: 0-8385-7701-6.

14. Ilfeld BM, Said ET, Finneran IV JJ, et al. Ultrasound-guided percutaneous peripheral nerve stimulation: neuromodulation of the femoral nerve for postoperative analgesia following ambulatory anterior cruciate ligament reconstruction: a proof of concept study. Neuromodulation. 2019;22:621–629.

15. Juckett L, Saffari TM, Ormseth B, Senger JL, Moore AM. The effect of electrical stimulation on nerve regeneration following peripheral nerve injury. Biomolecules. 2022;12(12):1856. doi:10.3390/biom12121856. PMID: 36551285; PMCID: PMC9775635

16. Schäfer B, Bahm J, Beier JP. Nerve Transfers Using a Dedicated Microsurgical Robotic System. Plast Reconstr Surg Glob Open. 2023 Aug 14;11(8):e5192. doi: 10.1097/GOX.0000000000005192. PMID: 37583397; PMCID: PMC10424892.

17. OpenAI. (2024). Figure created using ChatGPT

18. Manchikanti L, Sanapati MR, Soin A, Kaye AD, Kaye AM, Solanki DR, Chen GH, Nampiaparampil D, Knezevic NN, Christo P, Bautista A, Karri J, Shah S, Helm Ii S, Navani A, Wargo BW, Gharibo CG, Rosenblum D, Luthra K, Patel KG, Javed S, Reuland W, Gupta M, Abd-Elsayed A, Limerick G, Pasupuleti R, Schwartz G, Chung M, Slavin KV, Pampati V, Hirsch JA. Comprehensive Evidence-Based Guidelines for Implantable Peripheral Nerve Stimulation (PNS) in the Management of Chronic Pain: From the American Society Of Interventional Pain Physicians (ASIPP). Pain Physician. 2024 Nov;27(S9):S115–S191. PMID: 39565237.

19. Hardy PB, Wang BY, Chan KM, Webber CA, Senger JB. The use of electrical stimulation to enhance recovery following peripheral nerve injury. Muscle Nerve. 2024 Dec;70(6):1151-1162. doi: 10.1002/mus.28262. Epub 2024 Sep 30. PMID: 39347555.

